# First-trimester nocturnal cardiorespiratory coupling during home sleep testing and later pregnancy complications: a secondary analysis of the nuMoM2b cohort

**DOI:** 10.64898/2026.11.02.26367117

**Authors:** Yanqi Wu, Sima Asvadi, Myrthe van der Ven, M. Beatrijs van der Hout-van der Jagt, Elisabetta Peri, Rik Vullings, Judith OEH van Laar, Pedro Fonseca, Sebastiaan Overeem, Massimo Mischi, Xi Long

## Abstract

We tested whether direction-specific nocturnal cardiorespiratory coupling (CRC) in the first trimester differed in pregnancies that later developed obstetric complications. The analysis included 2,504 nulliparous women from the nuMoM2b cohort with valid level 3 home sleep recordings at 6–13 weeks’ gestation. Bidirectional CRC between RR intervals and abdominal respiratory effort was quantified using bivariate phase-rectified signal averaging in consecutive 5-min segments. Coupling strength, local transition dynamics, and waveform irregularity were summarized by nightly mean and standard deviation. Associations with gestational diabetes mellitus (GDM; n = 111), preeclampsia (n = 149), preterm birth (n = 192), and fetal macrosomia (n = 186) were examined using proportional-odds models with CRC feature quintile as the ordered outcome. Models included all four complications simultaneously and adjusted for maternal age, pre-pregnancy body mass index, race and ethnicity, gestational age at recording, relevant medical and family history, medication use, and apnea-hypopnea index >5 events·h^−1^. Holm correction was applied across outcomes. GDM was associated with lower respiration-to-heart rate coupling strength (cumulative odds ratio, 0.45; 95% CI, 0.31–0.64; P < 0.001) and shallower slope at the anchor point (0.63; 0.45–0.89; P = 0.03). Fetal macrosomia was associated with lower within-night variability in heart rate-to-respiration waveform irregularity (0.59; 0.45–0.77; P < 0.001). No associations with preeclampsia or preterm birth remained significant after correction. Direction-specific differences in nocturnal CRC were detectable in the first trimester in pregnancies that later developed GDM or resulted in fetal macrosomia.

**New & Noteworthy:** Using home sleep recordings from 2,504 women in the first trimester, we found direction-specific differences in maternal cardiorespiratory coupling in pregnancies that later developed gestational diabetes or resulted in fetal macrosomia. Gestational diabetes was associated with lower respiration-to-heart rate coupling strength, whereas fetal macrosomia was associated with lower within-night variability in heart rate-to-respiration coupling irregularity. No significant associations were observed for preeclampsia or preterm birth.

## 1. Introduction

Pregnancy complications such as gestational diabetes mellitus (GDM), preeclampsia (PE), preterm birth (PTB), and fetal macrosomia remain common and contribute substantially to maternal and perinatal morbidity worldwide [1–5]. These complications span distinct pathophysiological domains: GDM and fetal macrosomia share a common metabolic substrate centered on insulin resistance and altered glucose–lipid metabolism [6,7]. PE is strongly associated with placental and vascular dysfunction, whereas PTB has heterogeneous causes [8,9]. Because physiological maladaptation may begin before clinical diagnosis, identifying early changes in maternal regulation could help determine when differences associated with pregnancy complications first become detectable [10–13].

Much of the existing physiological literature on early pregnancy has focused on unidimensional autonomic markers, such as heart rate variability, which provide a broad picture of maternal autonomic adaptation but do not capture direction-specific coordination between cardiovascular and respiratory regulation [14–19]. Cardiorespiratory coupling (CRC) offers a way to characterize this interaction more directly. Previous studies have reported altered cardiorespiratory coupling in pregnancies complicated by PE, but these studies were generally small, clinic-based, cross-sectional, or conducted after the complication had already emerged [20-22]. As a result, it remains unclear whether altered CRC is already detectable in early pregnancy and whether any associations are shared across, or differ between, later obstetric complications. Sleep provides a relatively stable physiological context for assessing CRC because voluntary activity and behavioral variability are reduced. Home sleep recordings also allow prolonged assessment of cardiac and respiratory signals under unsupervised conditions [23,24]. In this study, we examined whether direction-specific nocturnal CRC, assessed from first-trimester home sleep recordings, was associated with subsequent GDM, PE, PTB, and fetal macrosomia. We further assessed whether the associations were shared across complications or were outcome specific.

## 2. Methods

### 2.1 Study design, participants, and data source

This study was a secondary analysis of the Nulliparous Pregnancy Outcomes Study: Monitoring Mothers-to-Be (nuMoM2b), a prospective multicenter cohort conducted at eight US clinical centers [25]. In the parent cohort, nulliparous women with singleton pregnancies were enrolled and attended study visits at 6–13 weeks’ gestation (visit 1), 16–21 weeks (visit 2), 22–29 weeks (visit 3), and at delivery, with prospective ascertainment of obstetric outcomes [25].

The present analysis used data from the nuMoM2b sleep ancillary study, in which a subset of participants underwent level-3 home sleep testing at visit 1 [26]. Participants were eligible for the current analysis if they had valid visit-1 home sleep recordings with analyzable electrocardiogram and respiratory effort signals, complete outcome ascertainment, and complete prespecified baseline covariates. Participants with pregestational diabetes were excluded because GDM was an outcome of interest. The cohort selection process is summarized in Supplemental Figure S1. Prespecified maternal covariates included age, pre-pregnancy body-mass index (BMI), race and ethnicity, polycystic ovary syndrome (PCOS), family history of diabetes or obstetric complications, psychotropic or cardiovascular medication use before or in early pregnancy, and gestational age at the sleep test. Family-history variables, although ascertained after visit 1 in the parent study, were treated as baseline variables because they reflect pre-pregnancy status. Respiratory events were scored centrally by certified scorers in the nuMoM2b sleep ancillary study; full scoring and quality-control procedures have been described previously [26]. Because the level-3 recordings did not include electroencephalography, the apnea–hypopnea index (AHI) was calculated as the number of apneas and hypopneas per hour of estimated sleep, with the sleep period estimated from recorded times in bed together with heart rate, respiratory, movement, and position signals. Hypopneas were defined as a reduction in nasal pressure (or the respiratory sum channel when nasal pressure was unavailable) lasting at least 10 seconds and accompanied by at least a 3% oxygen desaturation. AHI greater than 5 events per hour was included as a prespecified covariate in the primary adjusted models.

The Eunice Kennedy Shriver National Institute of Child Health and Human Development review board and the Eindhoven University of Technology ethical review board approved this secondary analysis. In the parent nuMoM2b study, written informed consent was obtained from all participants by trained study personnel at each site, as previously described [25].

### 2.2 Signal extraction and preprocessing

From visit-1 home sleep recordings, we used the single-lead bipolar electrocardiogram (ECG; 200 Hz), abdominal respiratory effort signal (50 Hz), and body activity signal derived from the internal X– Y gravity sensor (10 Hz) [26]. Because gross motion and sensor displacement can distort physiological signals in home recordings, segments with non-zero motion counts derived from the body activity signal were excluded [27].

For the electrocardiogram, the signal was band-pass filtered (0.5–40 Hz) and normalized to facilitate R-peak detection [28]. R-peaks were detected using HeartPy, which applies adaptive-threshold peak detection to the filtered signal [28], and consecutive RR intervals were calculated. RR intervals considered physiologically implausible or likely artifactual (>2.0 s, <0.4 s, or differing by more than 20% from the preceding interval) were excluded. The remaining RR intervals were retained as a beat-to-beat tachogram for subsequent BPRSA analysis.

For the respiratory effort signal, a low-pass filter (0.6 Hz) was applied to emphasize breathing-cycle morphology for peak and trough detection [29]. The signal was then centered by subtracting its median value. Respiratory peaks and troughs were identified from turning points based on sign changes in the first derivative. Detected respiratory events were excluded if they failed prespecified interval or amplitude thresholds [29].

### 2.3 Direction-specific CRC analysis

We quantified direction-specific CRC using bivariate phase-rectified signal averaging (BPRSA) [26]. BPRSA permits direction-specific analysis of nonstationary physiological signals by defining anchor points in a trigger signal and quantifying the corresponding response in a target signal [30]. For clarity, we use the term “heart rate” when describing the coupling direction, although the analysis was performed on the beat-to-beat RR-interval tachogram. We examined two distinct directional pathways. The first, respiration-to-heart rate coupling, used the onsets of exhalation and inhalation as respiratory anchor points. For each anchor point, a window of 2L+1 consecutive RR intervals (L=150 RR intervals) was extracted from the beat-to-beat tachogram, aligned by RR-interval lag, and averaged to construct a BPRSA waveform. For heart rate-to-respiration coupling, the respiratory effort signal was resampled to 10 Hz so that the 301-point window spanned a duration similar to that used in previous work [16]. Heart-rate acceleration and deceleration events served as cardiac anchor points. For each anchor point, 301 respiratory samples were extracted, aligned, normalized, and averaged to generate a BPRSA waveform. From each BPRSA waveform, we extracted maximum BPRSA amplitude (MBA), slope at the anchor point (SAP), and sample entropy (Entropy), representing coupling strength, local transition dynamics, and waveform irregularity or complexity, respectively. For sample entropy, the embedding dimension was set to 4 and the tolerance to 0.2 times the waveform standard deviation [16].

To characterize overnight dynamics, the trigger signal was divided into non-overlapping 5-minute segments. For each segment and each coupling direction, anchor points identified within that segment were used to construct a local BPRSA waveform; extraction windows in the target signal were allowed to extend beyond segment boundaries when needed. MBA, SAP, and Entropy were then computed for each local waveform. For each feature and coupling direction, nightly mean and nightly standard deviation were calculated, yielding 12 CRC features per participant (3 features × 2 summaries × 2 directions). For presentation, the same feature labels are used for both directions, with the coupling direction specified separately in the Results and Tables.

### 2.4 Ascertainment of pregnancy outcomes

Prespecified primary outcomes were GDM, PE, PTB, and fetal macrosomia. Outcome definitions followed nuMoM2b criteria and established obstetric definitions [25,31,32]. PTB was defined as delivery between 20 and 36 completed weeks of gestation. GDM was diagnosed using routine oral glucose tolerance testing at 24–28 weeks according to nuMoM2b prespecified criteria [25]. PE was defined as new-onset hypertension after 20 weeks with proteinuria or severe end-organ dysfunction [25]. Fetal macrosomia was defined as birthweight greater than 4000 g [31,32].

### 2.5 Statistical analysis

We assessed associations between first-trimester CRC features and subsequent outcomes using proportional-odds ordinal logistic regression. Because CRC features are dimensionless and were not assumed to relate linearly to obstetric risk, each feature was divided into cohort-specific quintiles and modeled as an ordered outcome.

For each feature and coupling direction, GDM, PE, PTB and fetal macrosomia were entered simultaneously as predictors, together with prespecified maternal covariates. Primary models adjusted for age, pre-pregnancy body-mass index, race and ethnicity, polycystic ovary syndrome, family history of diabetes or obstetric complications, psychotropic or cardiovascular medication use before or in early pregnancy, gestational age at the sleep test, and AHI greater than 5 events per hour. Results are reported as cumulative odds ratios with 95% confidence intervals. An odds ratio greater than 1 indicates a shift toward higher feature quintiles, whereas an odds ratio less than 1 indicates a shift towards lower feature quintiles.

Nightly mean features were prespecified as primary CRC outcomes, and nightly standard deviation features were considered secondary outcomes. Holm correction was applied to the four obstetric predictors within each model. For associations meeting Holm-adjusted p<0.05, we derived model-based predicted probabilities across feature quintiles and estimated 95% confidence intervals using 1000 bootstrap resamples.

Because the prespecified CRC features represented distinct aspects of coupling strength, transition dynamics, waveform irregularity, direction, and overnight variability, each feature was analyzed separately. Within each model, the four pregnancy complications constituted one family of hypotheses and were corrected using the Holm procedure. As a sensitivity analysis, the Benjamini–Hochberg procedure was applied jointly across all features, directions, summaries, and complications. All signal processing, BPRSA feature extraction, and statistical analyses were performed in Python 3.8.12.

### 2.6 Exploratory prediction analyses

To complement the association analyses, we examined whether first-trimester CRC features provided information about subsequent complications beyond routinely available maternal characteristics. For each complication, we compared a baseline model containing the prespecified maternal covariates (section 2.5) with an augmented model that additionally included the coupling features, and assessed whether adding coupling features improved the identification of affected women. Model performance was summarized by discrimination (area under the receiver operating characteristic curve and area under the precision–recall curve), with calibration and reclassification reported in Supplemental Figure S2. Model performance and its incremental change were estimated by bootstrap resampling (1000 resamples). These analyses were exploratory and were not intended to develop or validate a clinical prediction model.

## 3. Results

### 3.1 Cohort characteristics

Of the women enrolled in the nuMoM2b sleep ancillary study, 2504 met the eligibility criteria for the present analysis, having valid first-trimester electrocardiogram and respiratory effort signals, complete outcome ascertainment, and prespecified covariates. The mean gestational age at the home sleep recording was 12.2 weeks (SD 1.5). Overall, 205 (8.2%) women reported psychotropic medication use before or in early pregnancy, and 80 (3.2%) had an apnea–hypopnea index (AHI) greater than 5 events per hour. During follow-up, 111 (4.4%) women developed gestational diabetes mellitus (GDM), 149 (5.9%) developed preeclampsia (PE), 192 (7.7%) had preterm birth (PTB), and 186 (7.4%) delivered a macrosomic infant. Baseline characteristics and outcome frequencies are summarized in Table 1. When stratified by complication status (Supplemental Tables S1-S2), women who later developed GDM were on average older (29.9 vs 26.6 years), had a higher pre-pregnancy BMI (29.5 vs 26.3 kg/m^2^), and were more likely to have a family history of diabetes (31.5% vs 20.4%); a similar metabolic profile was seen for fetal macrosomia. By contrast, the PE and PTB groups differed primarily in the distribution of race/ethnicity and corresponding family histories.

**Table 1:** Maternal sociodemographic factors and pregnancy complications.

| **Characteristics** |  |
|---|---|
| Age, mean (SD), y | 26.8 (5.6) |
| Pre-pregnancy BMI, mean (SD), kg/m<sup>2</sup> | 26.3 (6.3) |
| Family history of diabetes, No. (%) | 522 (20.8%) |
| Family history of PE, No. (%) | 262 (10.4%) |
| Family history of PTB, No. (%) | 252 (10.4%) |
| PCOS, No. (%) | 140 (5.6%) |
| Race and Ethnicity, No. (%) |  |
| - Non-Hispanic White | 1552 (62.0%) |
| - Non-Hispanic Black | 289 (11.5%) |
| - Hispanic | 454 (18.1%) |
| - AAPI | 97 (3.9%) |
| - Other/Unknown | 113 (4.5%) |
| R-R Interval, mean (SD), ms | 847.8 (67.7) |
| Breathing Interval, mean (SD), s | 3.7 (0.4) |
| Use of cardiovascular medications, No. (%) | 122 (4.8%) |
| Use of psychotropic medications, No. (%) | 205 (8.2%) |
| AHI > 5, No. (%) | 80 (3.2%) |
| Gestational age, mean (SD), weeks | 12.2 (1.5) |
| **Pregnancy complications** |  |
| PTB | 192 (7.7%) |
| GDM | 111 (4.4%) |
| PE | 149 (5.9%) |
| Fetal macrosomia | 186 (7.4%) |

*Abbreviations: SD, standard deviation; y, years; BMI, body mass index; kg/m<sup>2</sup>, kilograms per square meter; No. (%), number (percentage); PCOS, polycystic ovary syndrome; AAPI, Asian American and Pacific Islander; AHI, apnea–hypopnea index; PTB, preterm birth; GDM, gestational diabetes mellitus; PE, preeclampsia*.

A comparison of baseline characteristics between the analytic cohort and eligible women not included in the analysis showed no statistically significant differences in age, BMI, medication use, or family history (Supplemental Table S3). The prevalence of all four pregnancy outcomes was also similar between groups (all p>0.05). Modest differences were observed in the distribution of two race/ethnicity categories, although absolute differences were small (<2 percentage points) and race/ethnicity was included as a covariate in all adjusted models. PCOS prevalence differed nominally, but this comparison was limited by a high proportion of missing PCOS data (>80%) in the parent cohort.

### 3.2 Primary associations: Nightly mean coupling features

After covariate adjustment and Holm correction, significant associations among the nightly mean features were observed only for GDM and only in the respiration-to-heart rate direction. Effect estimates for overall PE, PTB, and fetal macrosomia were generally close to the null (Figure 1). Specifically, women who later developed GDM were less likely to occupy higher quintiles of respiration-to-heart rate coupling strength (maximum BPRSA amplitude [MBA] nightly mean: cumulative odds ratio 0.45, 95% CI 0.31–0.64; Holm-adjusted p<0.001) and of local transition dynamics (slope at the anchor point [SAP] nightly mean: 0.63, 0.45–0.89; Holm-adjusted p=0.03). In the heart rate-to-respiration direction, cumulative odds ratios for nightly mean features were close to 1 across all outcomes.

**Figure 1:**
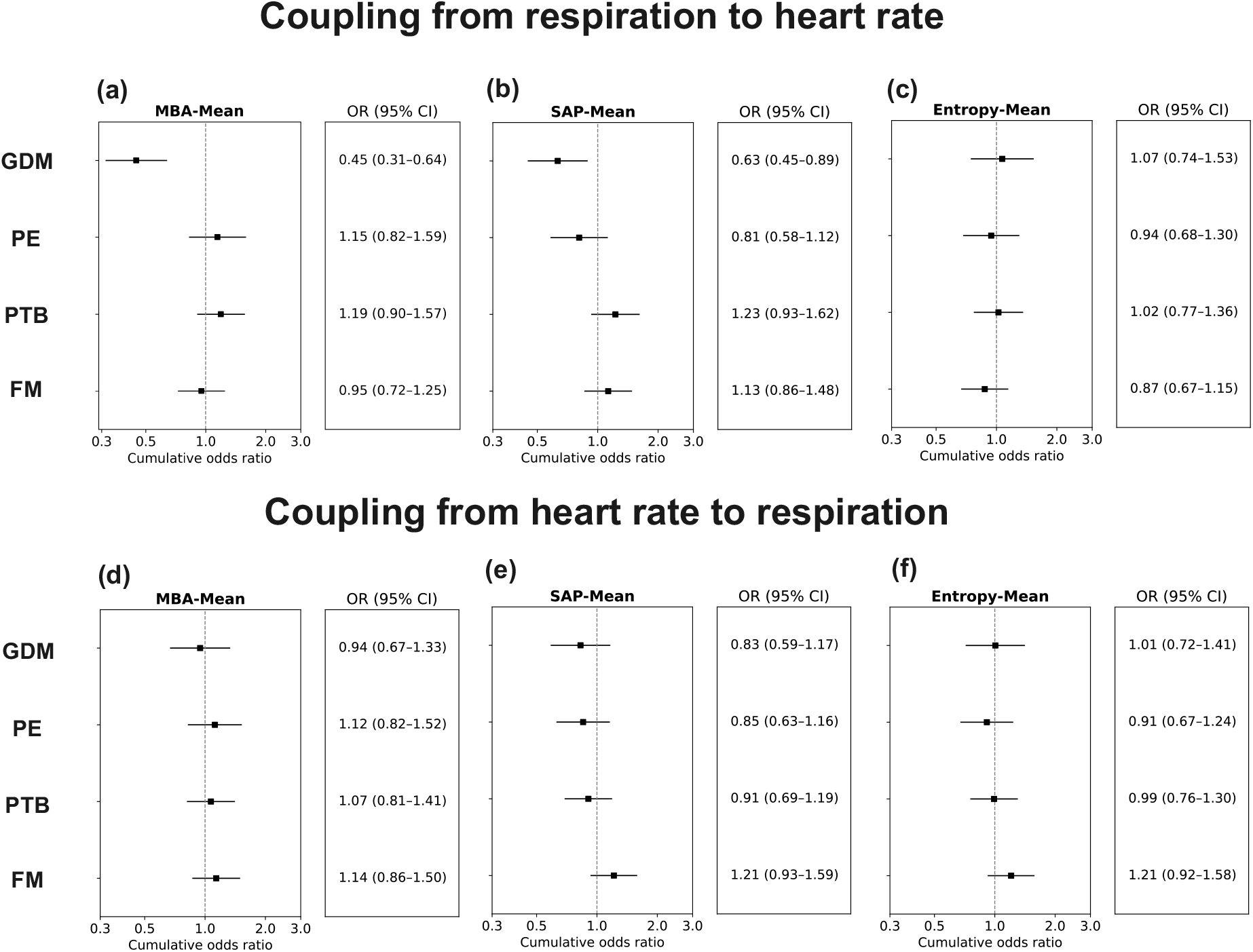
Associations of nightly mean CRC features with later pregnancy complications. Forest plots show cumulative odds ratios (ORs) and 95% CIs for cohort-specific quintiles of nightly mean features in the respiration-to-heart rate direction (A–C) and the heart rate-to-respiration direction (D–F). Dashed vertical lines indicate OR=1. Models were adjusted as described in Methods. MBA=maximum BPRSA amplitude. SAP=slope at the anchor point.

Model-based predicted probabilities illustrated the magnitude of this shift (Figure 2). For respiration-to-heart rate coupling strength, women who subsequently developed GDM had a higher predicted probability of falling into the lowest quintile than those who did not (33% vs 19%) and a lower probability of occupying the highest quintile (10% vs 20%). The corresponding differences in predicted probability were +14 percentage points for the lowest quintile and −10 percentage points for the highest quintile.

**Figure 2:**
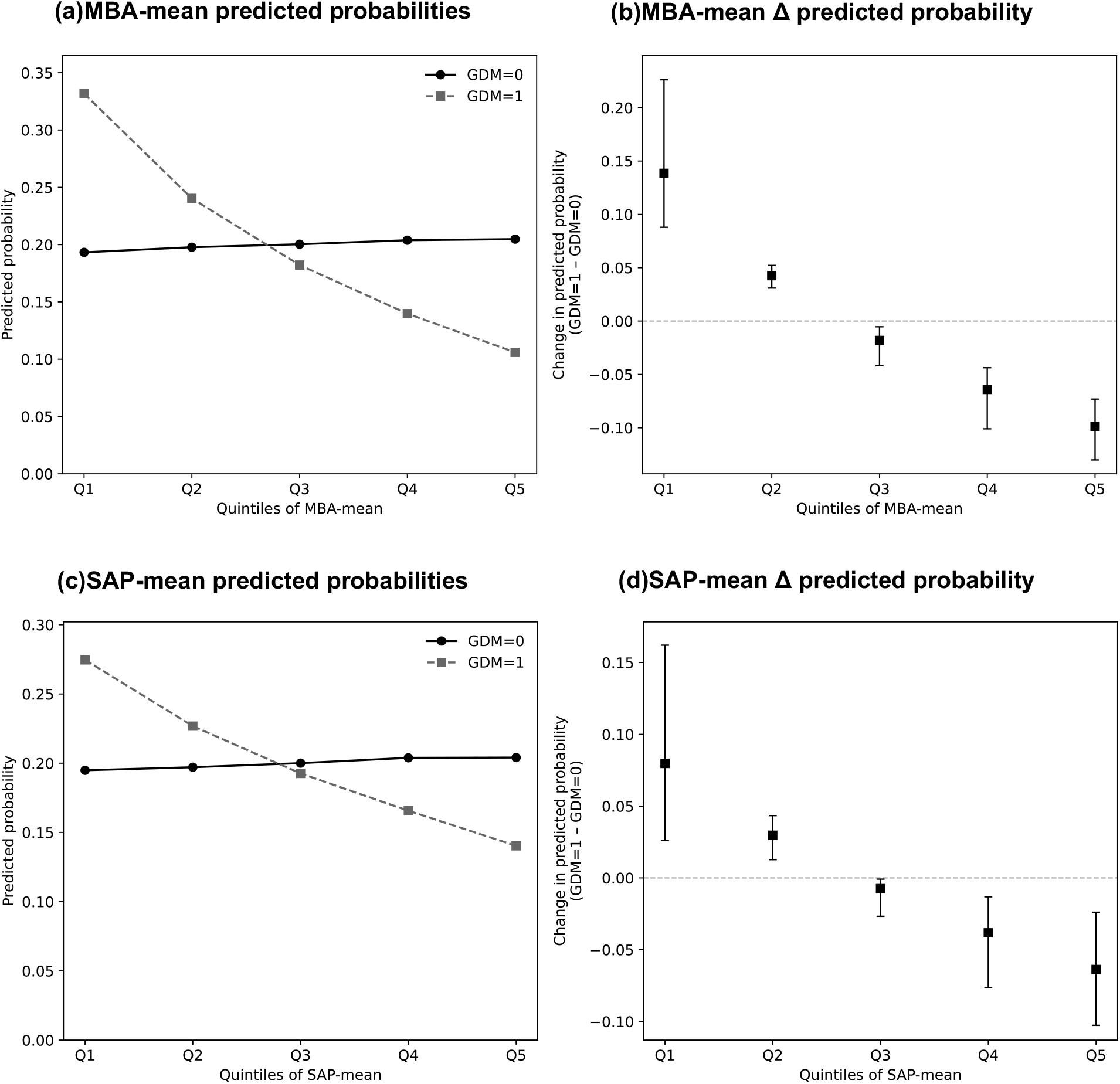
Model-based predicted probabilities across nightly mean CRC quintiles in women with and without gestational diabetes mellitus. Panels show predicted probabilities of each quintile and differences in predicted probability for respiration-to-heart rate MBA-mean (A, B) and SAP-mean (C, D). Probabilities were derived from the fitted proportional-odds models described in Methods 2.5. Error bars indicate 95% CIs from bootstrap resampling. MBA=maximum BPRSA amplitude. SAP=slope at the anchor point.

### 3.3 Secondary associations: Within-night coupling variability

Among nightly SD features, fewer associations remained significant after Holm correction than among nightly mean features. The clearest association was observed for fetal macrosomia in the heart rate-to-respiration direction (Figure 3).

**Figure 3:**
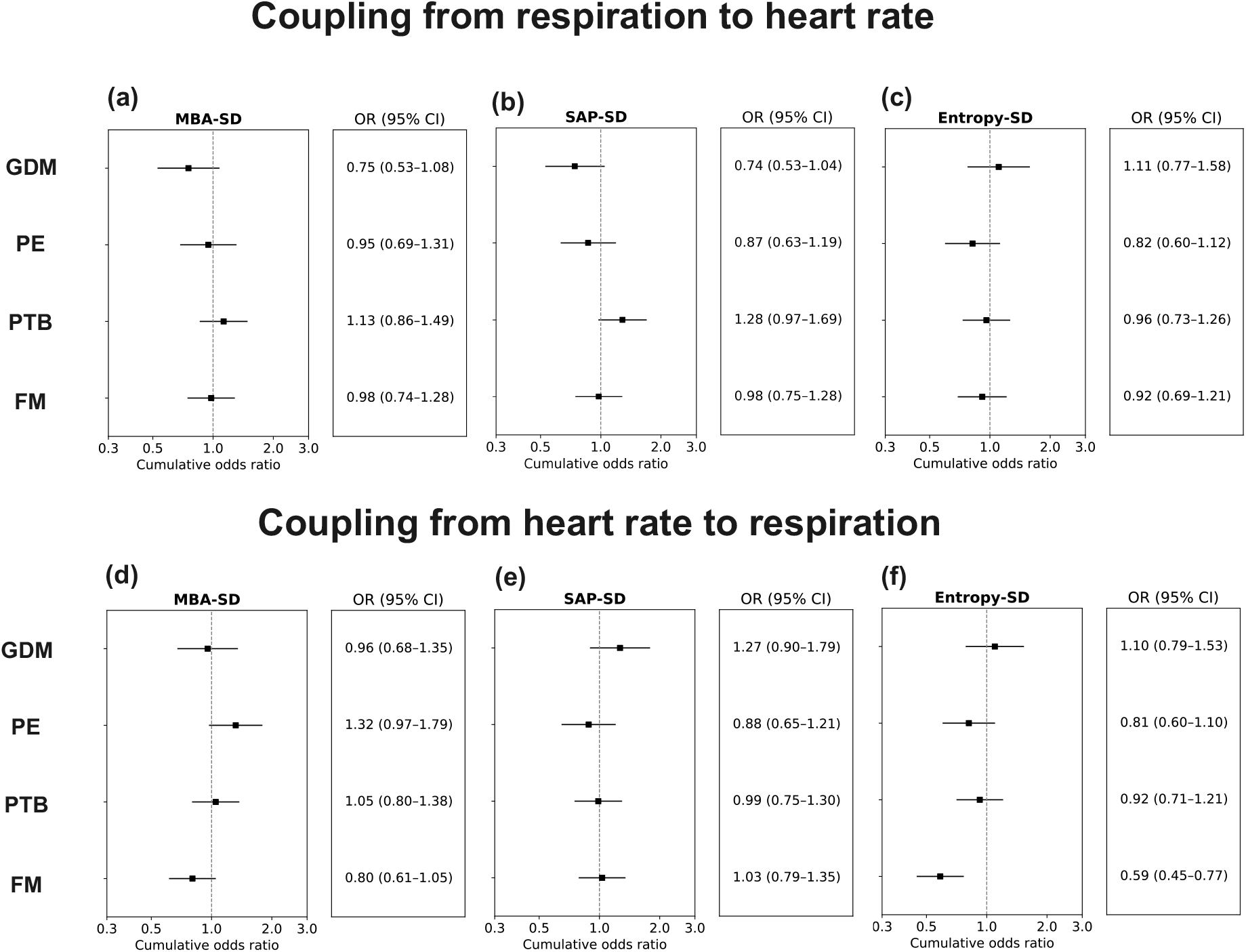
Associations of nightly variability in CRC features with later pregnancy complications. Forest plots show cumulative ORs and 95% CIs for cohort-specific quintiles of nightly standard deviation features in the respiration-to-heart rate direction (A–C) and the heart rate-to-respiration direction (D–F). Dashed vertical lines indicate OR=1. Models were adjusted as described in Methods 2.5. MBA=maximum BPRSA amplitude. SAP=slope at the anchor point.

Lower within-night variability in heart rate-to-respiration waveform irregularity (entropy nightly SD) was associated with fetal macrosomia (cumulative odds ratio 0.59, 95% CI 0.45–0.77; Holm-adjusted p<0.001). Women who later delivered a macrosomic infant were more likely to fall into the lowest entropy-SD quintile and less likely to fall into the highest quintile than women without macrosomia; the corresponding differences in predicted probability +10 and −7 percentage points, respectively (Figure 4). Variability features in the respiration-to-heart rate direction were not significantly associated with any prespecified outcome after Holm correction.

**Figure 4:**
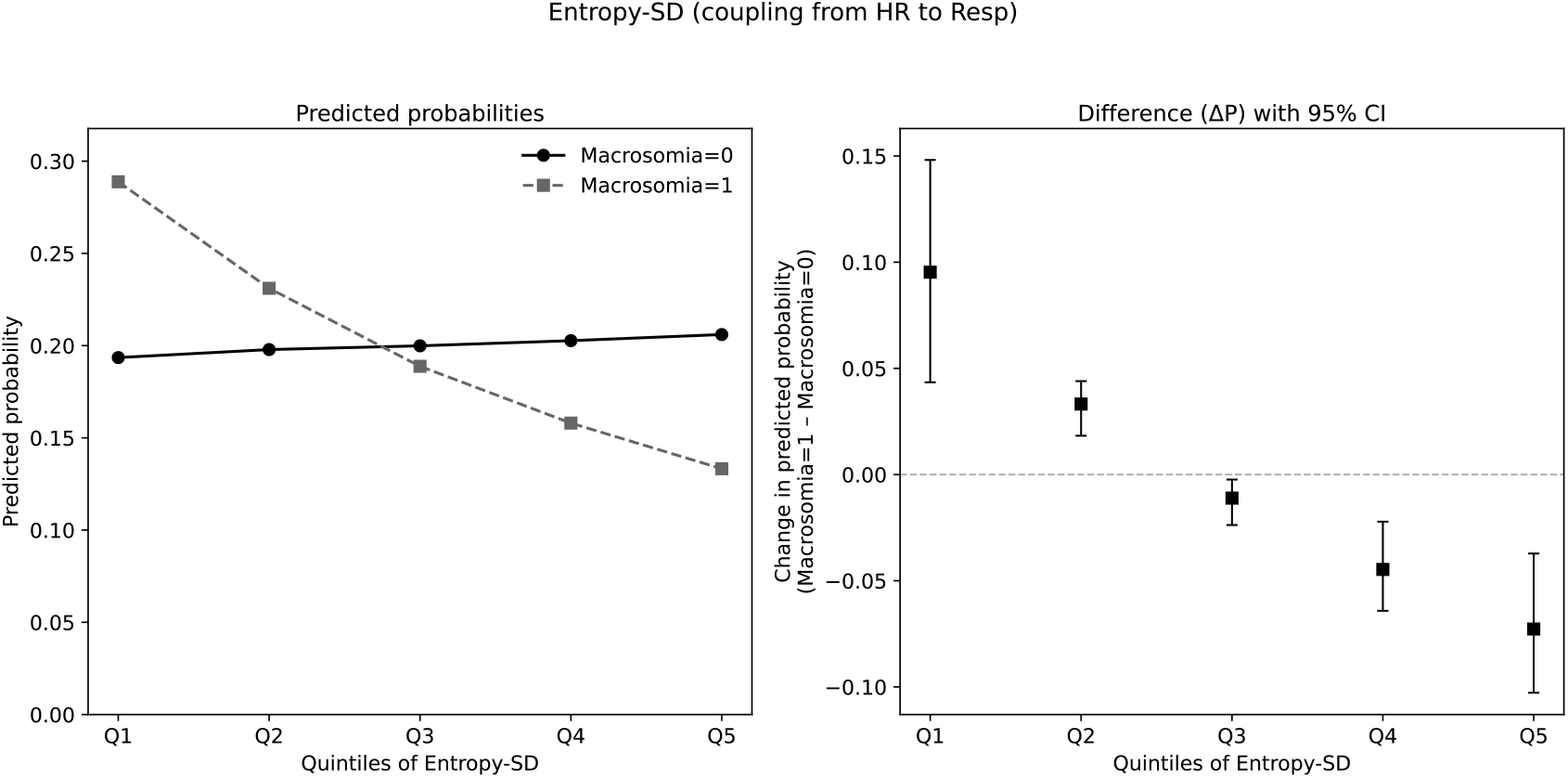
Model-based predicted probabilities across nightly variability quintiles in women with and without fetal macrosomia. Panels show predicted probabilities of each quintile and differences in predicted probability for heart rate-to-respiration Entropy-SD. Probabilities were derived from the fitted proportional-odds model described in Methods 2.5. Error bars indicate 95% CIs from bootstrap resampling. SD=standard deviation.

### 3.4 Exploratory prediction analyses

Across all four complications, the additional information from coupling features was most consistent for GDM, for which discrimination improved (area under the receiver operating characteristic curve 0.68 to 0.71). A smaller improvement in the same direction was seen for fetal macrosomia (0.62 to 0.64), whereas gains for the PTB and PE were minimal and inconsistent. The exploratory prediction results were consistent with the primary analyses, with the clearest incremental information observed for GDM and, to a lesser extent, fetal macrosomia.

## 4. Discussion

### 4.1 Main findings

In this secondary analysis of first-trimester home sleep recordings from 2,504 women, nocturnal CRC showed outcome- and direction-specific associations with later pregnancy complications. Women who later developed GDM had lower respiration-to-heart rate coupling strength and slower local transition dynamics, whereas fetal macrosomia was associated with lower within-night variability in heart rate-to-respiration coupling irregularity. Associations with PE and PTB were not significant after multiplicity correction.

### 4.2 Interpretation and relation to previous work

The GDM association was graded across CRC quintiles and was observed specifically in the respiration-to-heart rate direction. This is consistent with previous findings of reduced maternal autonomic modulation before GDM diagnosis [33]. Because CRC was measured at 6–13 weeks and GDM was diagnosed at 24–28 weeks, the observed coupling differences preceded clinical diagnosis. These differences may involve pathways related to insulin resistance and inflammatory signaling that affect central cardiorespiratory control [33,34,36], although the present study cannot establish the mechanism.

The pattern for fetal macrosomia was distinct from that of GDM and should be interpreted more cautiously, as it was supported by a single coupling feature rather than by convergent associations across features. Lower within-night variability in heart rate-to-respiration coupling irregularity may reflect reduced flexibility of cardiorespiratory regulation in pregnancies resulting in fetal macrosomia [15,35,36]. GDM and fetal macrosomia both involve altered maternal metabolic regulation, although the present data do not establish a shared mechanism.

The principal GDM and macrosomia findings remained significant after global false-discovery-rate correction. Similar associations were not observed for PE or PTB.

### 4.3 Physiological and translational implications

Nocturnal CRC may provide information about maternal physiological regulation in early pregnancy, but the modest improvements in the exploratory prediction analyses do not support its use as a standalone clinical predictor. Further evaluation will require independent cohorts. Future studies should determine whether the observed CRC associations differ across sleep stages rather than relying only on whole-night averages. External validation and prospective studies are also needed to assess their reproducibility and physiological relevance. The derivation of CRC features from unsupervised home recordings may facilitate their evaluation beyond the sleep laboratory. Future wearable systems capable of recording cardiac and respiratory signals during sleep could provide opportunities to evaluate these physiological features in larger prospective cohorts.

### 4.4 Strengths and limitations

The strengths of this study include its basis in a large prospective multicenter cohort of nulliparous women, the use of scalable first-trimester home sleep recordings, and a direction-specific analytical framework applied to synchronized ECG and respiratory effort signals. Evaluating multiple major outcomes within a single modeling framework allowed direct comparison of coupling associations across complications.

Several limitations should be considered. First, the level-3 home recordings did not include EEG-based sleep staging; because coupling varies by sleep state [16], averaging across the night may have diluted state-specific associations. Second, only a subset of the parent cohort had analyzable first-trimester recordings and complete data, which may limit generalizability. The analytic cohort and the remaining eligible women were similar in age, BMI, clinical characteristics, and the prevalence of all four pregnancy outcomes (Supplemental Table S3), although selection bias cannot be excluded. Third, adjustment for an AHI greater than 5 events per hour did not materially change the main estimates, although residual confounding by other dimensions of sleep-disordered breathing remains possible. Snoring was not explicitly detected during preprocessing. Although the RR-interval and respiratory-event quality criteria may have removed some obvious snoring-related disturbances, residual interference with the CRC estimates cannot be excluded. Fourth, by design, our coupling features emphasized dynamic interaction rather than absolute heart-rate or respiratory variability; the latter has also been linked to adverse outcomes [33,37] and could provide complementary information. Fifth, discretizing continuous coupling features into quintiles improved interpretability but may have reduced statistical power and obscured non-monotonic associations. Sixth, although we corrected for multiple testing within each model and confirmed the primary associations under a global false-discovery-rate correction, the observed association with macrosomia was confined to a single coupling feature and therefore requires independent replication. Finally, the prediction analyses were internally derived using bootstrap resampling, without an external validation cohort, and should be regarded as hypothesis-generating.

## 5. Conclusion

First-trimester nocturnal CRC showed direction- and outcome-specific associations with later GDM and fetal macrosomia, whereas associations with PE and PTB were not significant after multiplicity correction. These findings suggest that differences in maternal cardiorespiratory regulation were detectable in the first trimester in pregnancies that later developed GDM or resulted in fetal macrosomia. External validation and studies incorporating sleep-state information are needed to clarify the physiological basis and reproducibility of these associations.

## Data Availability

The data that support the findings of this study are available from the Eunice Kennedy Shriver National Institute of Child Health and Human Development Data and Specimen Hub upon application and approval. Restrictions apply to the availability of these data. Further information is available from the corresponding author upon reasonable request.

## Data availability

No new participant data were collected for this secondary analysis. Deidentified clinical and pregnancy-outcome data from the Nulliparous Pregnancy Outcomes Study: Monitoring Mothers-to-Be (nuMoM2b) are available through the Eunice Kennedy Shriver National Institute of Child Health and Human Development Data and Specimen Hub (NICHD DASH), subject to the repository’s application procedures and data-use agreement. Deidentified home sleep recordings are available through the National Sleep Research Resource, subject to its access requirements and data-use agreement. Data dictionaries and study documentation are available through the respective repositories. The authors are not permitted to redistribute the source participant-level data.

Analysis code used to generate the reported results will be available from the corresponding author upon reasonable request, subject to the terms of the original data-use agreements.

