## Supplemental materials for "First-trimester nocturnal cardiorespiratory coupling during home sleep testing and later pregnancy complications: a secondary analysis of the nuMoM2b cohort"

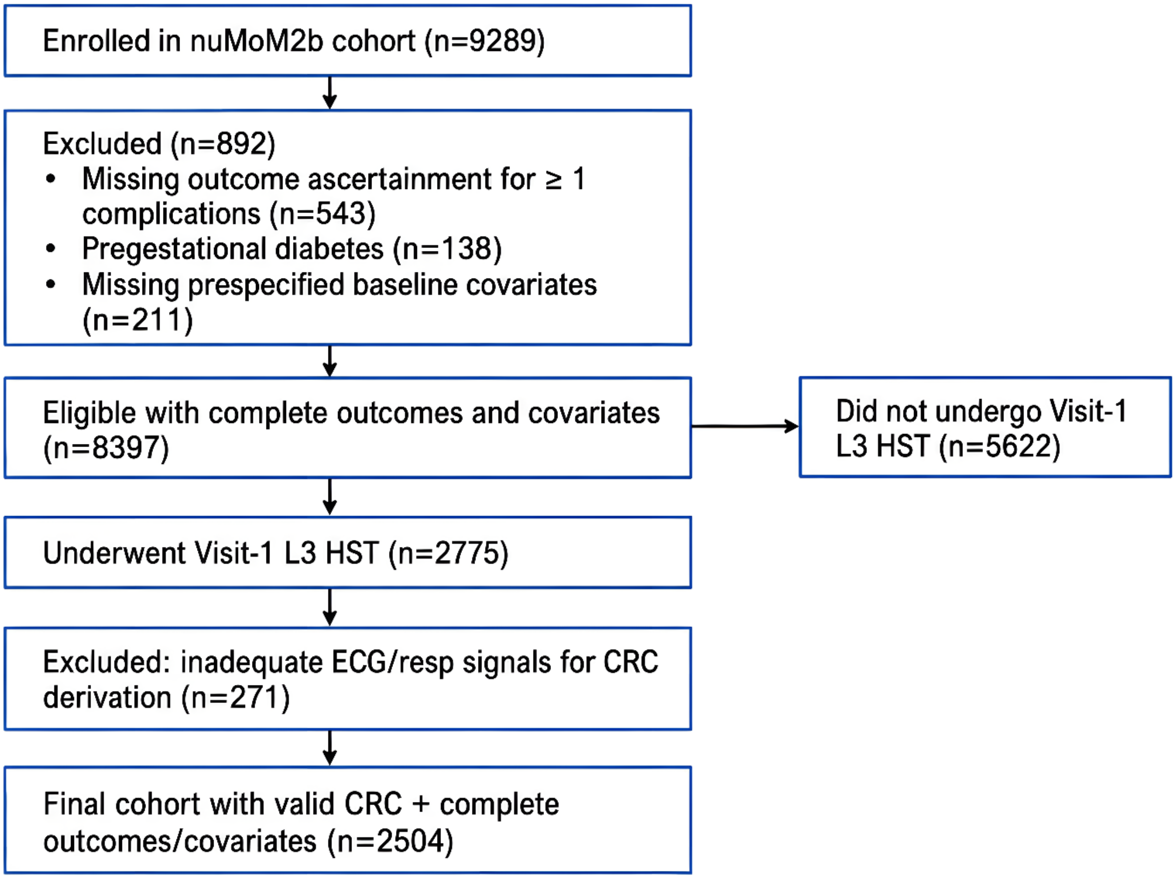


***Supplemental Figure S1: Selection of the first-trimester cardiorespiratory coupling analytic cohort from the nuMoM2b study.*** *Flow diagram showing derivation of the analytic cohort from the nuMoM2b cohort. Of 9289 enrolled participants, 892 were excluded for missing ascertainment of one or more of the prespecified obstetric outcomes (gestational diabetes mellitus, preeclampsia, preterm birth, or fetal macrosomia; n=543), pregestational diabetes (n=138), or missing prespecified baseline covariates (n=211), leaving 8397 eligible participants with complete outcomes and covariates. Of these, 2775 underwent a Visit-1 (6–13 weeks' gestation) level-3 home sleep test and 5622 did not. A further 271 participants were excluded because their electrocardiogram or respiratory effort signals were inadequate for cardiorespiratory coupling derivation, yielding a final analytic cohort of 2504 participants with valid cardiorespiratory coupling features and complete outcome and covariate data. HST=home sleep test. L3=level 3. ECG=electrocardiogram. resp=respiratory. CRC=cardiorespiratory coupling.*

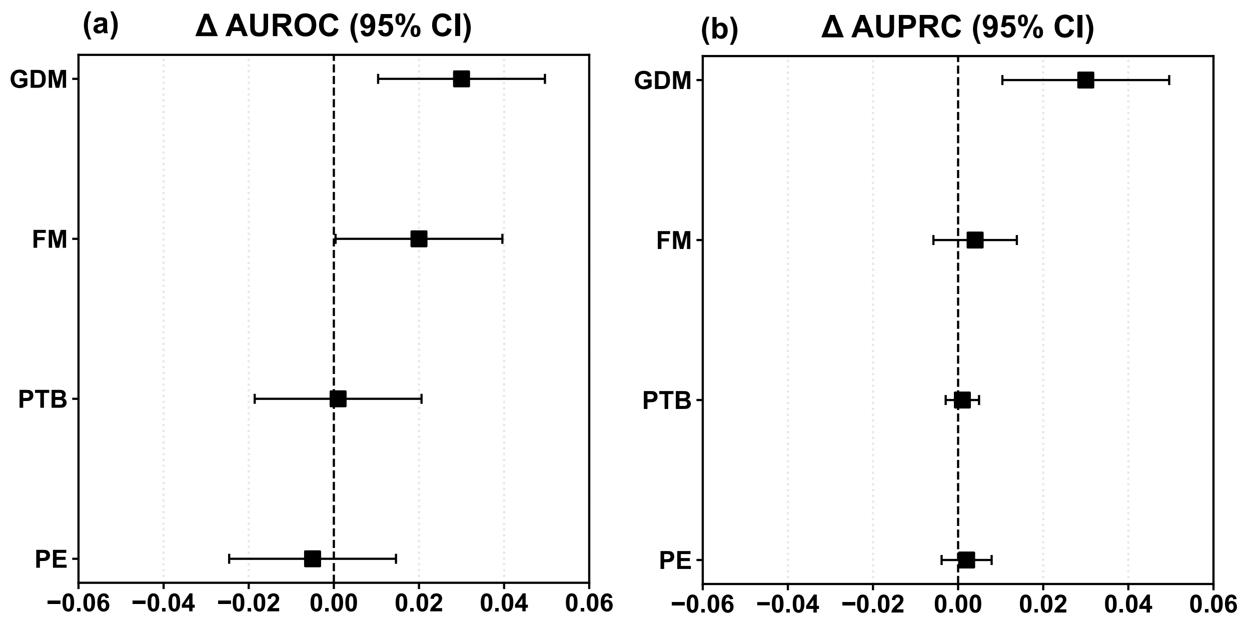


***Supplemental Figure S2: Incremental predictive performance after adding cardiorespiratory coupling features to baseline maternal characteristics.*** *Points show the mean change in AUROC and AUPRC by bootstrap resampling for GDM and fetal macrosomia. Error bars indicate 95% confidence range across resamples.*

***Supplemental Table S1. Baseline characteristics stratified by metabolic–growth outcomes***

| | **GDM** | | **Fetal macrosomia** | |
|---|---|---|---|---|
| | **Yes (n=108)** | **No (n=2396)** | **Yes (n=182)** | **No (n=2322)** |
| ***Demographics*** | | | | |
| Age, years | 29.9 (5.7) | 26.6 (5.5) | 27.7 (5.4) | 26.7 (5.6) |
| Pre-pregnancy BMI, kg/m$^2$ | 29.5 (7.4) | 26.2 (6.2) | 29.0 (7.0) | 26.1 (6.2) |
| ***Race/ethnicity*** | | | | |
| Non-Hispanic White | 65 (60.2%) | 1487 (62.2%) | 119 (65.4%) | 1433 (61.8%) |
| Non-Hispanic Black | 8 (7.4%) | 281 (11.7%) | 16 (8.8%) | 273 (11.8%) |
| Hispanic | 16 (14.8%) | 438 (18.3%) | 35 (19.2%) | 419 (18.1%) |
| Other* | 12 (11.1%) | 85 (3.6%) | 3 (1.6%) | 94 (4.1%) |
| ***Clinical characteristics*** | | | | |
| PCOS | 9 (8.3%) | 131 (5.5%) | 8 (4.4%) | 132 (5.7%) |
| Cardiovascular medication use | 7 (6.5%) | 115 (4.8%) | 12 (6.6%) | 110 (4.7%) |
| Psychotropic medication use | 14 (13.0%) | 193 (8.1%) | 14 (7.7%) | 193 (8.3%) |
| AHI, events/h | 2.4 (4.1) | 0.9 (2.0) | 1.2 (2.3) | 0.9 (2.2) |
| ***Family history*** | | | | |
| Diabetes | 34 (31.5%) | 488 (20.4%) | 37 (20.3%) | 485 (20.9%) |
| Preeclampsia | 12 (11.1%) | 250 (10.5%) | 15 (8.2%) | 247 (10.7%) |
| Preterm birth | 12 (11.1%) | 250 (10.5%) | 15 (8.2%) | 247 (10.7%) |

| | | | | |
|---|---|---|---|---|
| ***First-trimester physiological measurements*** | | | | |
| Gestational age at recording, weeks | 12.2(1.9) | 12.3 (1.5) | 12.0 (1.4) | 12.3 (1.5) |
| ***Outcome co-occurrence*** | | | | |
| GDM | 111 (100%) | 0 (0%) | 10 (5.5%) | 98 (4.2%) |
| Fetal macrosomia | 10 (9.3%) | 172 (7.2%) | 186 (100%) | 0 (0%) |
| Preeclampsia | 12 (11.1%) | 137 (5.7%) | 4 (2.2%) | 145 (6.3%) |
| Preterm birth | 17 (15.7%) | 176 (7.4%) | 3 (1.6%) | 190 (8.2%) |

Data are mean (SD) or n (%). *Includes American Indian, Asian, Native Hawaiian, and other. AHI=apnoea–hypopnoea index. BMI=body-mass index. GDM=gestational diabetes mellitus. PCOS=polycystic ovary syndrome.

***Supplemental Table S2. Baseline characteristics stratified by vascular–placental outcomes***

| | **Preeclampsia** | | **Preterm birth** | |
|---|---|---|---|---|
| | **Yes (n=149)** | **No (n=2355)** | **Yes (n=193)** | **No (n=2311)** |
| ***Demographics*** | | | | |
| Age, years | 26.3 (5.9) | 26.8 (5.5) | 27.2 (6.1) | 26.7 (5.5) |
| Pre-pregnancy BMI, $kg/m^2$ | 29.2 (7.0) | 26.2 (6.2) | 27.2 (7.0) | 26.3 (6.2) |
| ***Race/ethnicity*** | | | | |
| Non-Hispanic White | 78 (52.3%) | 1474 (62.7%) | 116 (60.1%) | 1436 (62.2%) |
| Non-Hispanic Black | 33 (22.1%) | 256 (10.9%) | 38 (19.7%) | 251 (10.9%) |
| Hispanic | 29 (19.5%) | 425 (18.1%) | 25 (13.0%) | 429 (18.6%) |
| Other* | 1 (0.7%) | 96 (4.1%) | 4 (2.1%) | 93 (4.0%) |
| ***Clinical characteristics*** | | | | |
| PCOS | 8 (5.4%) | 132 (5.6%) | 16 (8.3%) | 124 (5.4%) |
| Cardiovascular medication use | 8 (5.4%) | 114 (4.8%) | 12 (6.2%) | 110 (4.8%) |
| Psychotropic medication use | 10 (6.7%) | 197 (8.4%) | 13 (6.7%) | 194 (8.4%) |
| AHI, events/h | 1.6 (4.3) | 0.9 (2.0) | 0.8 (1.5) | 0.9 (2.2) |
| ***Family history*** | | | | |
| Diabetes | 40 (26.8%) | 482 (20.5%) | 39 (20.2%) | 483 (20.9%) |
| Preeclampsia | 27 (18.1%) | 235 (10.0%) | 30 (15.5%) | 232 (10.1%) |
| Preterm birth | 14 (9.4%) | 248 (10.5%) | 23 (11.9%) | 239 (10.4%) |
| ***First-trimester physiological measurements*** | | | | |
| Gestational age at recording, weeks | 12.5 (1.4) | 12.3 (1.5) | 12.5 (1.3) | 12.2 (1.5) |
| ***Outcome co-occurrence*** | | | | |
| Preeclampsia | 149 (100%) | 0 (0%) | 49 (25.4%) | 100 (4.3%) |

| | | | | |
|---|---|---|---|---|
| Preterm birth | 49 (32.9%) | 144 (6.1%) | 192 (100%) | 0 (0%) |
| GDM | 12 (8.1%) | 96 (4.1%) | 17 (8.8%) | 91 (3.9%) |
| Fetal macrosomia | 4 (2.7%) | 178 (7.6%) | 3 (1.6%) | 179 (7.8%) |

Data are mean (SD) or n (%). *Includes American Indian, Asian, Native Hawaiian, and other. AHI=apnoea–hypopnoea index. BMI=body-mass index. GDM=gestational diabetes mellitus. PCOS=polycystic ovary syndrome.

***Supplemental* Table S3. Comparison of baseline characteristics between the analytic cohort and eligible women not included in the analysis**

| Characteristic | Analytic cohort (n=2504) | Not included (n=5893) | p value |
|---|---|---|---|
| **Demographics** | | | |
| Age, years | 26.8 (5.6) | 27.0 (5.7) | 0.069 |
| Pre-pregnancy BMI, $kg/m^2$ | 26.3 (6.3) | 26.2 (6.2) | 0.578 |
| **Race/ethnicity** | | | |
| Non-Hispanic White | 1524 (62.2%) | 3647 (61.5%) | 0.550 |
| Non-Hispanic Black | 287 (11.7%) | 797 (13.4%) | 0.032 |
| Hispanic | 436 (17.8%) | 940 (15.8%) | 0.029 |
| Other* | 96 (3.9%) | 272 (4.6%) | 0.174 |
| **Clinical characteristics** | | | |
| PCOS† | 134 (5.5%) | 220 (3.7%) | <0.001 |
| Cardiovascular medication use | 121 (4.9%) | 315 (5.3%) | 0.484 |
| Psychotropic medication use | 203 (8.3%) | 445 (7.5%) | 0.223 |
| **Family history** | | | |
| Diabetes | 511 (20.8%) | 1243 (21.0%) | 0.914 |
| Preeclampsia | 257 (10.5%) | 593 (10.0%) | 0.500 |
| Preterm birth | 256 (10.4%) | 606 (10.2%) | 0.754 |
| **Pregnancy outcomes** | | | |
| GDM | 107 (4.4%) | 258 (4.3%) | 0.974 |
| Preeclampsia | 149 (6.1%) | 362 (6.1%) | 0.855 |
| Preterm birth | 192 (7.9%) | 482 (8.1%) | 0.567 |
| Fetal macrosomia | 186 (7.4%) | 450 (7.6%) | 0.609 |

*Data are mean (SD) or n (%). P values are from Welch t-tests (continuous variables) or $\chi^2$ tests (categorical variables) and are provided for descriptive comparison only, without adjustment for multiple testing. The analytic cohort comprises women with valid first-trimester cardiorespiratory coupling features, complete outcome ascertainment, and prespecified covariates. *Includes American Indian, Asian, Native Hawaiian, and other. †PCOS data had a high proportion of missing values (~80%) in the parent cohort; the apparent difference should be interpreted with caution. AHI=apnoea–hypopnoea index. BMI=body-mass index. GDM=gestational diabetes mellitus. PCOS=polycystic ovary syndrome.*
